# Effectiveness of Physical Activity Interventions Utilizing Wearables and Smartphone Applications for Individuals with Cardiovascular Diseases and Stroke: A Systematic Review and Meta-analysis

**DOI:** 10.1101/2025.11.05.25339609

**Authors:** Ajith Kumar Vemuri, Yasaman Rahimi, Alireza Vafaei Sadr, Seyyed Sina Hejazian, Shouhao Zhou, Jonathan Hakun, Christopher Sciamanna, Vida Abedi, Ramin Zand

## Abstract

**Background:** Lack of physical activity is a major risk factor for Cardiovascular diseases (CVDs). Wearables and smartphones offer potential advantages in cost, accessibility, and scale to change physical activity This systematic review aims to determine if interventions comprising smartphones and wearables are effective in enhancing physical activity among individuals with CVD.

**Methods:** PubMed, Cochrane, and Web of Science databases were searched for RCTs, published since January 2000, on wearables and smartphone applications to enhance physical activity among CVD patients. Non-randomized trials, interventions without a smartphone/wearable component were excluded. Risk of bias was assessed by the Cochrane Collaboration tool. Meta-analyses were performed to assess the pooled effect on steps per day, distance, VO2, and (moderate-to-vigorous) physical activity in minutes per day.

**Results:** Fourteen RCTs were included. Interventions comprising smartphones and wearables resulted in a mean difference of 1097.4 (95% CI = [409.2-1785.6]; p=0.0018) steps per day, and 3.9 (95% CI = [0.2-7.6]; p=0.0413) minutes of moderate-vigorous physical activity per day compared to control groups.

**Conclusion:** This meta-analysis showed that interventions comprising smartphones and wearable devices are effective at increasing physical activity among CVD patients. Wearables and smartphones could provide accessible and tailored interventions to enhance physical activity.

## 1. Introduction

Recent evidence highlights a decade-long rise in CVD burden, the leading cause of mortality globally, especially in low and middle-income countries,^1–3^ underscoring the need to develop cost-effective and accessible interventions.^2,4^ Over the years, an abundance of research has firmly established a lack of physical activity as a major risk factor for CVD and the health benefits of light, moderate, and vigorous physical activity.^1,5–7^ Although effective physical activity interventions exist for CVD patients, their implementation is limited by high costs, potentially reaching $60,000 per Quality-Adjusted Life Year (QALY) gained.^8^

Smartphones and wearable trackers offer potential advantages in accessibility, cost, scale, and the ability to deliver interventions. Although smartphones and wearable trackers generally provide reliable measurements of physical-activity metrics (e.g., steps, heart rate) in cardiac populations, in certain subgroups with impaired gait (e.g., after stroke or when using assistive devices), measurement accuracy may decline and should therefore be interpreted with caution^9–13^ Additionally, the efficacy of smartphones and wearable trackers in improving physical activity remains unclear. Multiple studies have tried to address this question. Pfaeffli et al’s systematic review in 2016 addressed the use of mHealth interventions for CVD self-management, however, the review only included two studies on physical activity.^14^ In 2018, Coorey et al’s meta-analysis concluded that larger, controlled studies of longer duration are required to establish the effectiveness of smartphone applications for CVD self-management.^15^ Kacie et al’s 2021 systematic review and meta-analysis on smartphone applications’ effectiveness to improve physical activity in CVD patients provided initial evidence on this question.^16^ However, their study included individuals with hypertension in the CVD cohort and omitted trials utilizing wearable trackers.

This systematic review and meta-analysis aim to explore the recent evidence and determine Using the PICOTS framework, this review aimed to determine whether, among adults with cardiovascular diseases, digital interventions utlizing smartphones and wearable activity trackers, compared with usual care or attention controls, increase objectively measured physical activity outcomes, steps per day, minutes per day of moderate-to-vigorous physical activity, six-minute walk distance, or peak VO_2_ at the end of the intervention period in randomized controlled trials. Additionally, this study seeks to identify the behavioral theories and techniques used in these interventions.

## 2. Methods

### Search Strategy

This systematic review is reported according to the Preferred Reporting Items for Systematic Reviews and Meta-Analyses statement (PRISMA)^17^ and the Cochrane Handbook for Systematic Reviews of Interventions.^18^ A systematic literature search was conducted to identify relevant studies published from 2000 up to February 2025. The search encompassed three electronic databases: PubMed, the Cochrane Central Register of Controlled Trials (CENTRAL), and Web of Science. The search aimed to find randomized controlled trials (RCTs) evaluating the effectiveness of physical activity interventions using wearables or smartphone applications to promote physical activity in cardiovascular populations, compared to a control group. Search terms included relevant MeSH terms (e.g., ‘Mobile Applications’, ‘Wearable Devices’, ‘Exercise’, ‘Cardiovascular Diseases’, ‘Randomized Controlled Trial’) combined with keywords and text words searched in titles and abstracts. The initial search yielded 556 results from PubMed, 1536 from CENTRAL, and 625 from Web of Science. Additionally, the reference lists of included studies and relevant systematic reviews were manually screened to identify any potentially eligible trials not captured by the database search. The full detailed search strategies used for each database are available in the Appendix.

### Inclusion/Exclusion Criteria

Studies were included in this meta-review if they met specific criteria based on the Population, Intervention, Comparison, Outcome, and Study Design (PICOS)^19^ framework. Eligible studies were RCTs published in English between January 2000 and February 2025. The study population consisted of both adults (aged 18 years or older) and adolescents with established cardiovascular disease (such as coronary heart disease, stroke, or heart failure). Interventions were required to focus on promoting physical activity, utilizing wearable technology (e.g., activity trackers, smartwatches) and/or smartphone or tablet-based mobile health applications (mHealth apps) as a core component, either alone or as part of a multicomponent program. The comparison group could be any control condition, including usual care, attention control, waitlist, or an active control. Finally, studies needed to report at least one post-intervention measure of physical activity, assessed either objectively or via self-report.

Studies were excluded if they were non-randomized designs, published before 2000, or those without cardiovascular disease. Interventions relying solely on video conferencing or phone calls without a core wearable/smartphone application element were also excluded. Furthermore, studies that did not report physical activity outcomes were ineligible. Study protocols, conference abstracts, and other non-peer-reviewed publications were not included.

### Screening and Selection Process

The study selection process involved two stages (Figure 1). Initially, titles and abstracts retrieved from the database searches were independently screened by two review authors against the eligibility criteria. Subsequently, the full texts of potentially relevant articles were obtained and independently assessed for final inclusion by two review authors. Any disagreements encountered during the title/abstract screening or full-text review stages were resolved through discussion and consensus between the two reviewers. If consensus could not be reached, a third review author was consulted for a final decision.

**Figure 1.**
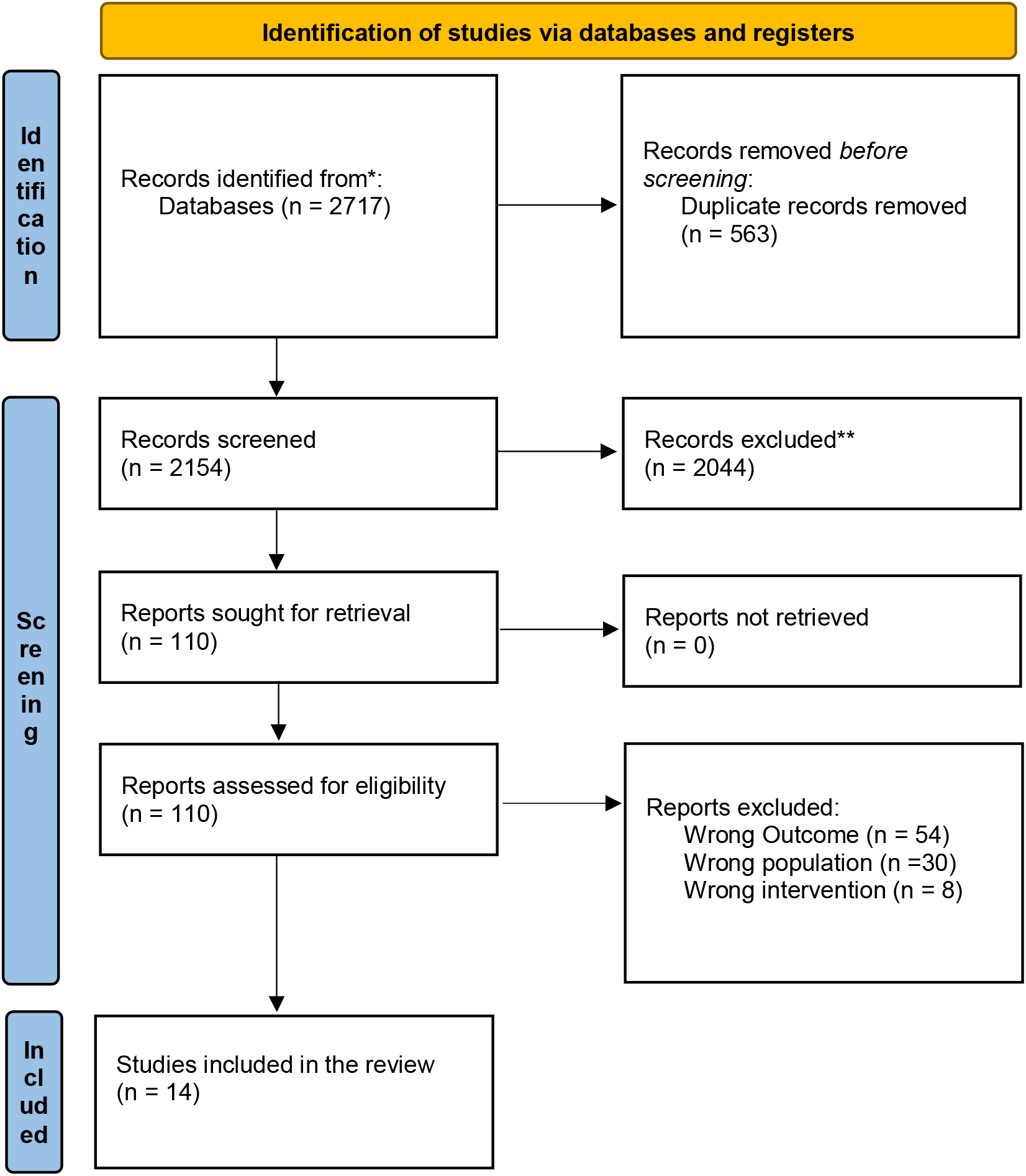
PRISMA flow diagram demonstrating the flow of studies through the review

**Figure 2.**
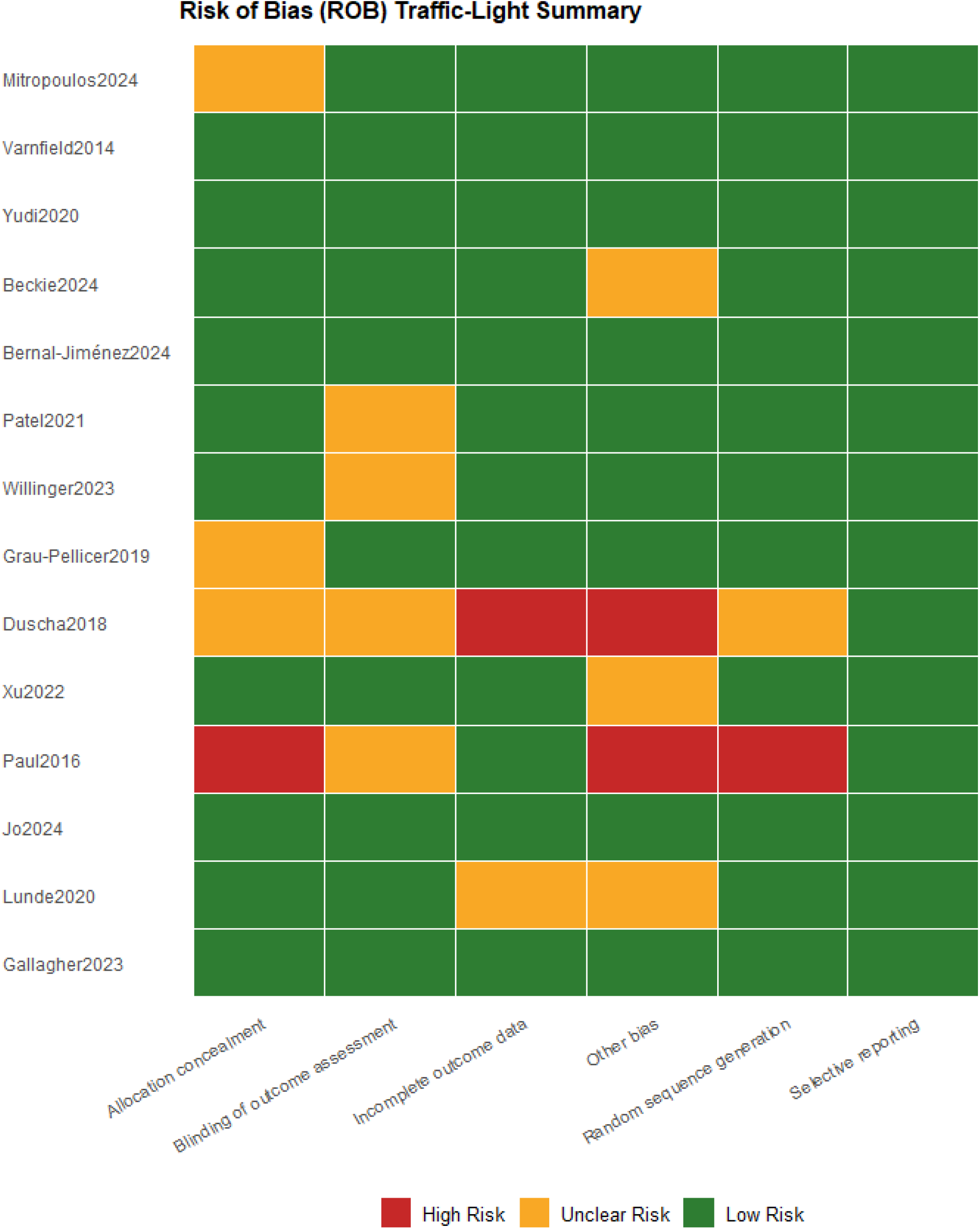
Bias plot of 14 studies in meta-analysis

**Figure 3.**
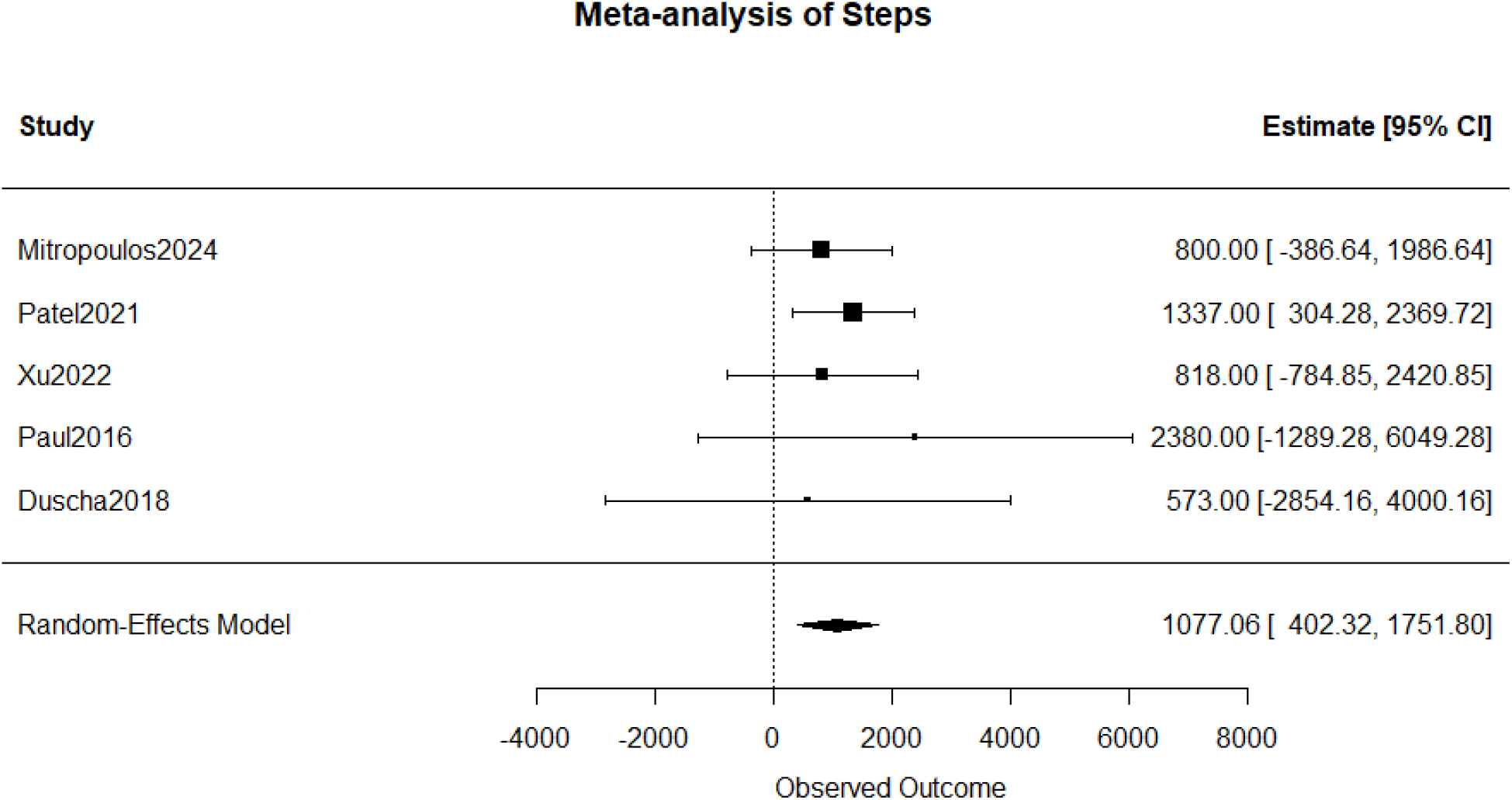
Meta-analysis of Steps per Day

**Figure 4.**
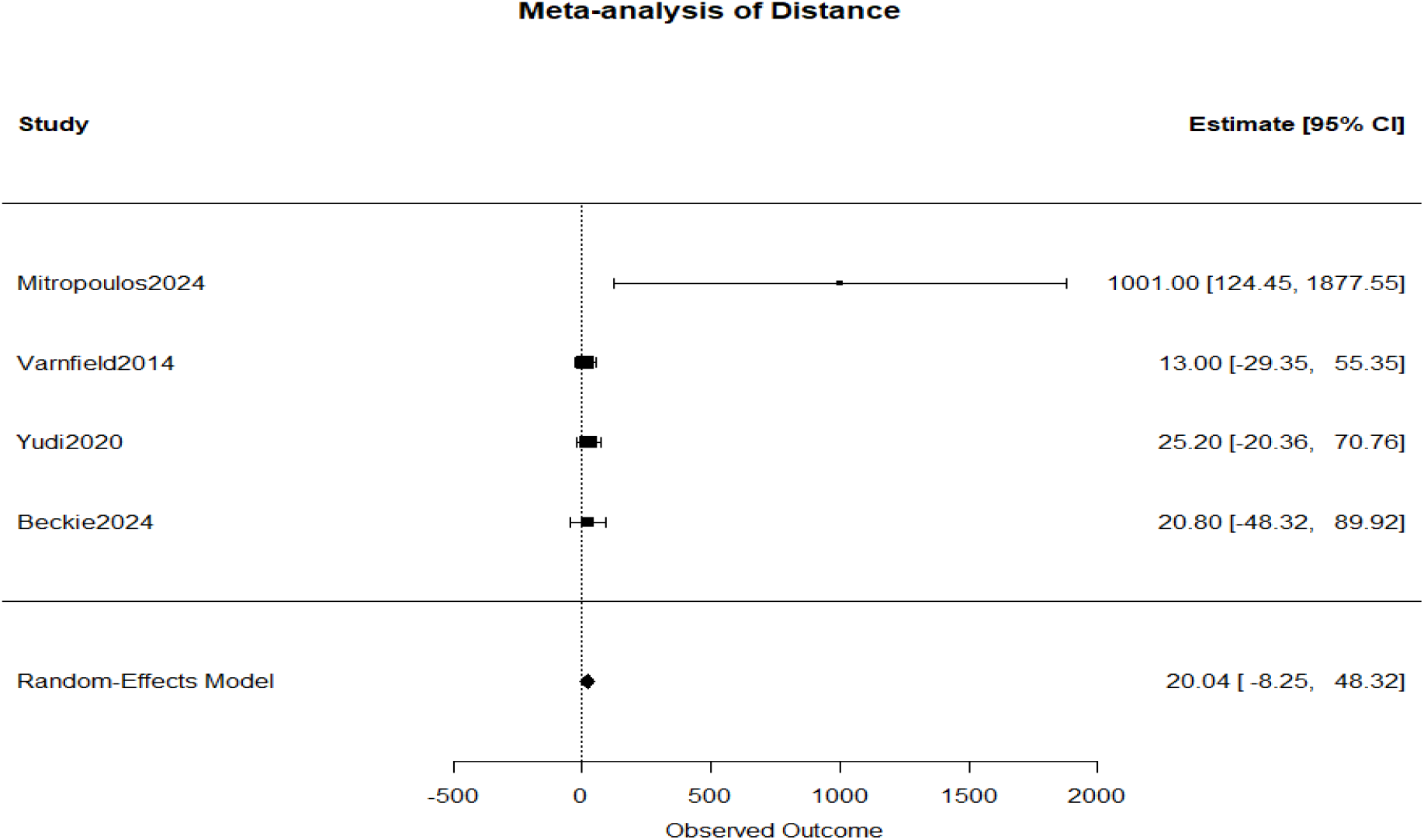
Meta-analysis of Distance

**Figure 5.**
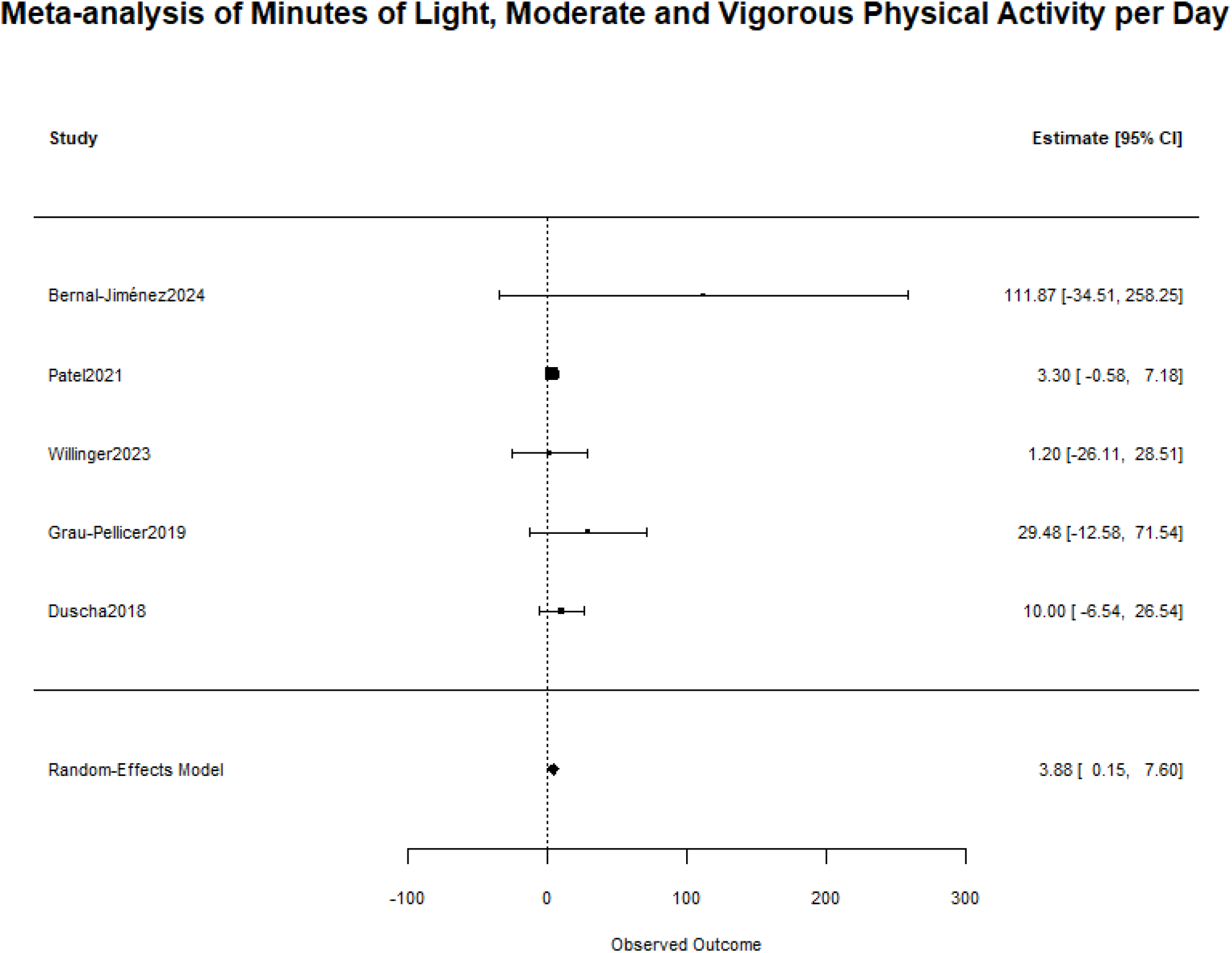
Meta-analysis of minutes in light, moderate, and vigorous physical activity

**Figure 6.**
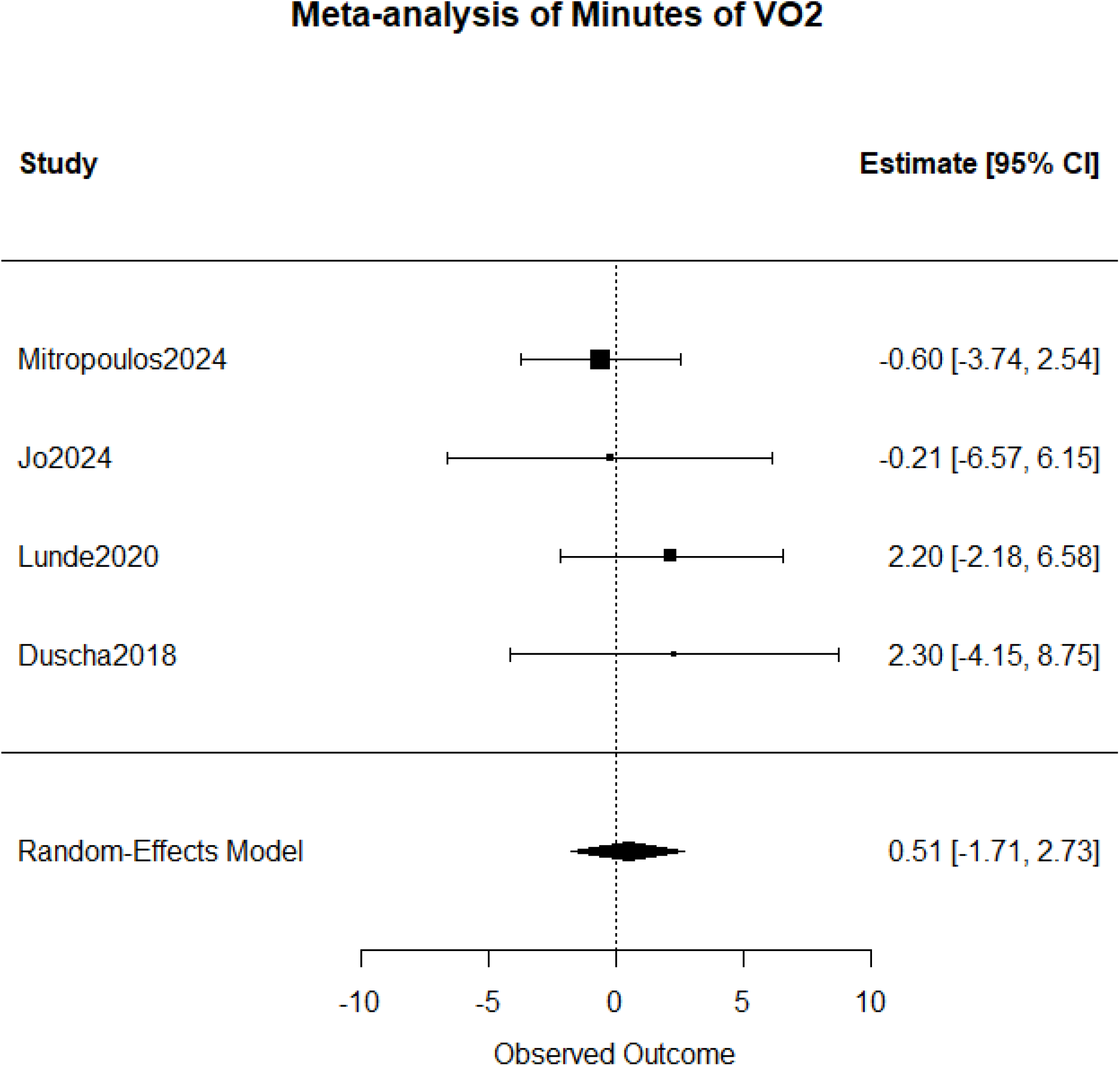
Meta-analysis of VO2

### Data Extraction

The extracted information covered several key areas. Study characteristics included the study identifier (e.g., first author and year), publication year, location, stated aims, trial type (RCT), and intervention duration. Population details encompassed sample size, baseline demographics, and the study’s inclusion and exclusion criteria. Detailed information on the intervention was extracted, including the type of wearable devices or smartphones used, the specific components and delivery mode of the intervention, and the nature of the control group comparison.

Outcome measures were documented, including primary and secondary outcomes reported by the study authors, specific physical activity metrics used (e.g., steps, MVPA), the timing of outcome measurements, and the study’s main conclusions regarding intervention effects. Furthermore, information on the behavioral theory explicitly cited by the authors was recorded, if available. The specific Behavior Change Techniques (BCTs) implemented within each intervention were identified and coded by the review authors based on intervention descriptions, using the BCT Taxonomy v1 (BCT-T v1).^20^ For studies featuring multiple relevant intervention arms, data were extracted for the arm that utilized the wearable and/or smartphone application with the most additional components compared to the control condition. If essential data were missing or unclear in the published report, supplementary materials were reviewed.

### Risk of Bias Assessment

The methodological quality and risk of bias for each included study were assessed independently by two review authors using the Cochrane risk of bias tool (RoB 2), which evaluates six key domains. Disagreements between the two reviewers regarding the risk of bias ratings were resolved through discussion and consensus, or by consulting a third review author if necessary. Each domain was judged as having a ‘Low risk’, ‘Unclear risk’, or ‘High risk’ of bias based on criteria adapted from the Cochrane Handbook.

The following domains were assessed. 1) Random sequence generation: This risk was judged based on whether the method used (e.g., computer-generated numbers) was appropriate to produce comparable groups. Studies not describing the method adequately were rated as unclear risk, while methods like sequential allocation were rated high risk. 2) Allocation concealment: Assessment focused on whether the method used (e.g., central randomization, sealed opaque envelopes) was sufficient to prevent foreknowledge of group assignment before or during participant enrolment. Studies not describing the concealment method were rated as unclear risk. 3) Blinding of outcome assessment: This was evaluated considering the methods used to measure the primary physical activity outcomes. Studies using primarily objective measures (e.g., accelerometers) assessed by blinded personnel were generally rated low risk. Studies relying heavily on subjective self-report outcomes without clear assessor blinding were considered at higher or unclear risk of detection bias. Blinding of participants and personnel delivering the interventions was noted as generally infeasible in these types of behavioral studies. 4) Incomplete outcome data: This domain assessed whether attrition and exclusions were adequately reported and addressed. Low risk was assigned if attrition was low (<15%) and balanced, reasons were reported, and appropriate analyses (e.g., intention-to-treat) were used. Higher or differential attrition led to unclear or high-risk ratings. 5) Selective reporting: Risk was assessed by comparing reported outcomes against those listed in trial registrations or protocols. Studies reporting outcomes as pre-specified were rated low risk. 6) Other bias: This domain considered other potential sources of bias, such as significant baseline imbalances between groups that were not adjusted for.

### Statistical Analyses

All meta-analyses were performed using random-effects models (DerSimonian–Laird and restricted maximum likelihood estimators) implemented in the “meta” and “metaphor” R packages. Each study-specific mean difference was treated as a random effect assumed to vary around an overall pooled mean difference, with between-study variance (τ^2^) estimated using restricted maximum likelihood (REML). This approach accounts for expected heterogeneity in interventions, devices, and populations and yields more conservative pooled estimates than fixed-effects models. The primary outcomes synthesized quantitatively were changes in physical activity levels, specifically steps per day, minutes of moderate-to-vigorous physical activity (MVPA) per day, distance [meters], and peak oxygen consumption (VO2 peak) [ml/kg/min].

Change-from-baseline scores (mean and standard deviation, SD) were calculated or extracted for the intervention and control groups for each study. Where necessary, data reported in weekly units were converted to daily averages (e.g., dividing minutes per week by 7). A random-effects model was employed for all meta-analyses to account for expected heterogeneity between studies. The overall treatment effect was calculated as the pooled mean difference (MD) with corresponding 95% confidence intervals (CIs) comparing the change in the intervention group to the change in the control group for each outcome.

Statistical heterogeneity across studies was quantified using the I^2^ statistic. Values of I^2^ were interpreted according to standard thresholds (<25% indicating low, 25-50% moderate, and >50% high heterogeneity). Visual inspection of forest plots also contributed to the assessment of heterogeneity. Potential publication bias was assessed by visual examination of funnel plot asymmetry for each outcome analysis. Formal testing for funnel plot asymmetry was conducted using Egger’s regression test.

## 3. Results

Title and abstract screening of 2717 unique articles led to the full-text assessment of 110 articles, with 14 studies^21–34^ ultimately included in this review (Fig. 1).

### 3.1 Population and Intervention Characteristics

The characteristics of the included studies and interventions involved 1057 participants with cardiovascular diseases from various populations. Coronary Heart Disease (CHD) or Myocardial Infarction (MI) was the most frequently studied population, encompassing 8 of the 14 studies included in this review. These studies investigated various specific populations within this category, including patients diagnosed with Myocardial Infarction (MI) or post-Acute Myocardial Infarction (post-AMI) (3 studies), patients broadly defined with CHD or Coronary Artery Disease (CAD) (3 studies), and individuals with CHD/Acute Coronary Syndrome (1 study). Additionally, specific subgroups were targeted, such as a cohort comprised exclusively of women with CHD (1 study). Two studies included participants with stroke. Additionally, two studies included patients with broader CVD diagnoses, and one study on adolescents (12-18 years) with moderate-to-complex congenital heart disease (Table 1). In all studies, physical activity was promoted through a smartphone or wearable device. In most studies, the application was supported by a pedometer or accelerometer.

Analysis of the 14 studies identified 27 unique Behavior Change Techniques (BCTs) utilized across the interventions, as shown in Table 2, with a total of 130 BCT instances recorded. The most frequently employed BCTs centered on monitoring and feedback, with ‘Self-monitoring of behavior’ present in all 14 studies and ‘Feedback on behavior’ in 13. Goal-setting techniques were also highly prevalent: ‘Goal setting (behavior)’ and ‘Review behavior goal(s)’ were each identified in 11 studies. ‘Instruction on how to perform a behavior’ was also very common, found in 10 studies. Other frequently used techniques included ‘Prompts/cues’ (9 studies), ‘Graded tasks’ (7 studies), and ‘Verbal persuasion about capability’ (7 studies). A broader range of BCTs addressing areas such as social support, other reward/threat techniques, natural consequences, and identity were employed less frequently across the included studies. Control groups differed across studies, varying from usual care to some form of education through a presentation, booklet, or education visit.

### Outcome Measures

Most studies measured physical activity objectively with an external accelerometer or pedometer, or with a smartphone’s built-in accelerometer or pedometer. One study subjectively measured physical activity using a questionnaire. Most studies reported their results in mean minutes of physical activity per day or daily step count. Other reported outcomes were mean hours of physical activity per week, mean minutes of physical activity per week, kilocalories per day, and metabolic equivalent of task (MET) per day. Eleven studies reported a change in physical activity levels between the baseline and the end of the intervention, and seven studies reported post-treatment physical activity levels.

### 3.2 Risk of Bias of Included Studies

The risk of bias assessment for the 14 included studies revealed variability in methodological quality across different domains. Regarding random sequence generation, the majority of studies (12 out of 14) were rated at low risk, typically employing computer-generated sequences often managed by independent personnel or services. One study was rated unclear risk due to insufficient description, and one pilot study utilizing sequential allocation was rated high risk. For allocation concealment, 8 studies achieved a low risk rating through methods like sealed opaque envelopes or central randomization. However, the risk was unclear in 5 studies due to inadequate reporting of the concealment mechanism, and one pilot study (Paul, 2016)^30^ using sequential allocation was rated high risk. Blinding of outcome assessment was rated low risk in 9 studies, which generally cited blinding of assessors and often used objective primary outcomes. The remaining 5 studies were rated unclear risk, because blinding status was not mentioned. The risk of bias from incomplete outcome data was judged low in 10 studies, characterized by low (<15%), balanced attrition and the use of intention-to-treat analyses. Three studies presented an unclear risk due to attrition rates around 15% or slight imbalances, while one study was rated high risk because of high (>20%) and differential attrition. The risk of selective reporting bias was deemed low for all 14 studies; trials were generally pre-registered, and reporting aligned with stated methods and outcomes. Concerns regarding other potential biases were common, with 7 studies rated as unclear risk, frequently arising from comparisons between technology-based interventions and less intensive usual care controls, raising possibilities of performance bias or Hawthorne effects. Three studies were assessed as high risk due to combined factors such as pilot study limitations, non-random allocation, or significant differential attrition.

### 3.3 Effect on Physical Activity

#### Effect on Steps

Four studies (k = 4)^21,26,28–30^ reported changes in daily steps from baseline to post-intervention. A total of 321 participants (166 in the intervention group and 155 in the control group) were included in these studies. Random effects models revealed a statistically significant improvement in steps in the intervention groups compared to the control group (mean difference = 1097.38, 95% CI = [409.17-1785.60]; z = 3.13, p = 0.0018). The analysis indicated no statistically significant heterogeneity among the included studies (I^2^ = 0.0%, 95% CI = [0.0%-84.7%]; Cochran’s Q = 1.03, df = 3, p = 0.7930).

#### Effect on Distance

This meta-analysis included four studies (k = 4),^21–24^ encompassing 334 participants (175 in the intervention group and 159 in the control group), that reported on changes in distance (meters). Three studies measured the distance as 6-minute walk tests, whereas one study reported the daily distance walked. The results from random effects models indicated no statistically significant difference in distance between the intervention and control groups (mean difference = 20.04, 95% CI = [-8.25-48.32]; z = 1.39, p = 0.1650). The heterogeneity analysis revealed moderate heterogeneity across the studies (I^2^ = 39.6%, 95% CI = [0.0%-79.5%]; Cochran’s Q = 4.97, df = 3, p = 0.1742).

#### Effect on Moderate and Vigorous Activity

The meta-analysis of light, moderate to vigorous physical activity included five studies (k = 5),^25–28,33^ comprising 480 participants (252 in the intervention group and 228 in the control group). The random effects models demonstrated a statistically significant improvement in moderate to vigorous physical activity in the intervention groups compared to the control group (mean difference = 3.88, 95% CI = [0.15-7.60]; z = 2.04, p = 0.0413). The heterogeneity analysis indicated low heterogeneity across the studies (I^2^ = 3.9%, 95% CI = [0.0%-80.0%]; Cochran’s Q = 4.16, df = 4, p = 0.3845).

#### Effect on VO2 (maximal oxygen consumption)

This meta-analysis of VO2 involved four studies (k = 4)^21,28,31,34^ with a total of 198 participants (99 in the intervention group and 99 in the control group). The results from random effects models showed no statistically significant difference in VO2 between the intervention and control groups (MD = 0.51, 95% CI = [-1.71-2.73]; z = 0.45, p = 0.6519). The heterogeneity analysis indicated no statistically significant heterogeneity across the studies (I^2^ = 0.0%, 95% CI = [0.0%-84.7%]; Cochran’s Q = 1.40, df = 3, p = 0.7063).

## 4. Discussion

This meta-analysis suggests that interventions comprising smartphones and wearable devices are effective at increasing physical activity among patients with cardiovascular disease (CVD). Specifically, both steps per day and minutes in moderate-to-vigorous physical activity significantly increased compared to controls. No significant effect was found in increasing VO2 and distance walked. ‘Self-monitoring of behavior’ and ‘Feedback on behavior’ were the most commonly used behavior techniques in the interventions. Only two interventions explicitly mentioned the underlying behavioral theories – ‘Phases of change theory’^ref^ and ‘Bandura’s social cognitive theory’^ref^ in their interventions.

A significant mean difference of 1097 steps per day was found between the intervention and control groups. This finding is comparable to previous meta-analyses measuring steps per day in various other chronic disease populations, including CVD.^16,35,36^ Although there is no established evidence of the clinical significance of this increase in steps per day among CVD patients, a similar increase in 350 to 1100 steps among obstructive pulmonary disease patients is reported to have resulted in a reduction of hospital admissions.^37^ In cardiac populations, however, the clinical relevance of such a change remains uncertain, particularly given that device accuracy can vary with gait characteristics and mobility status. Nevertheless, these results indicate that smartphone/wearable-based interventions show promise in promoting physical activity and may achieve greater benefits with continued advances in sensor accuracy and more comprehensive intervention designs that integrate behavioral-change strategies and personalized feedback.

A significant mean difference of 3.88 minutes per day in moderate to vigorous physical activity was found between intervention and control groups. This finding slightly differs from the existing meta-review on CVD patients, where the difference was 5.8 minutes per day.^16^

No significant increases were found in distance and VO2. We could not find any pool analysis measuring the change in distance and VO2 among CVD patients. This is due to the fact that most of the studies are recent (after 2020). Although the increase in distance measured in the 6m walk test (6MWT) is not significant, a 20m increase was observed between the intervention and control groups. There is evidence that small improvements in 6MWT negatively predict cardiovascular events.^38^

Analysis of the 14 studies identified 27 unique Behavior Change Techniques (BCTs) utilized across the interventions, as shown in Table 2, with a total of 130 BCT instances recorded. The most frequently employed BCTs centered on monitoring and feedback, with ‘Self-monitoring of behavior’ present in all 14 studies and ‘Feedback on behavior’ in 13. Goal-setting techniques were also highly prevalent: ‘Goal setting (behavior)’ and ‘Review behavior goal(s)’ were each identified in 11 studies. ‘Instruction on how to perform a behavior’ was also very common, found in 10 studies. Other frequently used techniques included ‘Prompts/cues’ (9 studies), ‘Graded tasks’ (7 studies), and ‘Verbal persuasion about capability’ (7 studies). A broader range of BCTs addressing areas such as social support, other reward/threat techniques, natural consequences, and identity were employed less frequently across the included studies. Control groups differed across studies, varying from usual care to some form of education through a presentation, booklet, or education visit.

‘Self-monitoring of behavior’ and ‘Feedback on behavior’ were highly prevalent in the interventions. These behavioral techniques are known to be associated with increased physical activity among CVD patients. However, the lack of details on how these interventions are designed and why certain components of the interventions are included, and the content of the intervention messages not being publicly available, restricts the replicability of these interventions. Additionally, action and coping planning,^39–41^ which are proven to have moderating effects on sustaining physical activity, are rarely present in the analyzed interventions. Utilization of these proven behavioral techniques, along with emerging techniques such as Just-in-Time-Adaptive interventions,^42–44^ could further help improve physical activity among individuals with CVD.

Our study has the following limitations: Most studies in this review focused on results directly post-intervention. Therefore, we could not obtain an insight into the sustainability of increased physical activity levels. Further research should include long-term follow-up assessments. Furthermore, step-count accuracy and reliability vary across devices and populations. Research indicates that wearable and smartphone step-count accuracy can decline at slow walking speeds or in individuals with atypical gait patterns, such as those recovering from stroke or using assistive devices. Accordingly, the pooled step-count increase observed in this meta-analysis should be interpreted with caution in such subgroups, as measurement error may attenuate or inflate true changes in physical activity. Also, it would be interesting to obtain more insight into adherence to the use of wearables and smartphone applications and factors influencing adherence, e.g., personal preferences for apps and behavior change techniques. Greater adherence may mediate the effect of the intervention on physical activity. With more information on sustainability, adherence, and long-term effectiveness, wearables and smartphone apps could be designed that are most effective in promoting physical activity to optimize impact on public health. In this review, it was not possible to study the effectiveness of each behavior change technique since they co-occur with other intervention characteristics and behavior change techniques. Lastly, the CVD population was predominantly comprised of heart disease patients, restricting generalization to other CVD patients, such as stroke survivors.

## Conclusion

Smartphone and wearable interventions were effective in increasing physical activity for those with CVD. The results indicate a potential benefit of smartphone apps in CVD patients, although caution is warranted due to the low number of trials and most being in coronary heart disease. Larger-scale research is needed to investigate the effect of smartphone and wearable applications across CVD diagnoses to be able to determine the ideal length and components of successful interventions.

## Data Availability

This a systematic review and meta analysis, all data is publicly available through published manuscripts

## Pubmed

(((Cell Phones[MeSH] OR Mobile Applications[MeSH] OR Actigraphy[MeSH] OR Wearable Devices[MeSH] OR mhealth[MeSH]) OR (smartphone*[tiab] OR wearable*[tiab] OR “activity monitor”[tiab] OR “activity tracker”[tiab] OR “mhealth”[tiab] OR “m-health”[tiab] OR “mobile health”[tiab] OR actigraph*[tiab] OR “wearable technolog*”[tiab] OR “wearable device*”[tiab] OR “smart band*”[tiab] OR “smart watch*”[tiab]))

AND

((Exercise[MeSH] OR Motor Activity[MeSH] OR Sedentary[MeSH] OR Physical Fitness[MeSH]) OR (“physical activity”[tiab] OR exercis*[tiab] OR “physical fitness”[tiab] OR “activity level*”[tiab] OR “exercise behavio*”[tiab] OR “physical exercise”[tiab] OR “sport”[tiab]))

AND

((Randomized Controlled Trial[pt] OR Clinical Trial, Randomized[MeSH]) OR (“randomized controlled trial”[tiab] OR “RCT”[tiab] OR “randomised controlled trial”[tiab]))

AND

((Cardiovascular Diseases[MeSH] OR Stroke[MeSH] OR Myocardial Infarction[MeSH] OR Heart Failure[MeSH] OR Cerebral Infarction[MeSH] OR Heart Diseases[MeSH]) OR (“CVD”[tiab] OR “Cardiovascular disease”[tiab] OR “Cardiovascular Disorder”[tiab] OR “Stroke”[tiab] OR “Brain Infarction”[tiab] OR “Heart Failure”[tiab] OR “heart disease”[tiab] OR “MI”[tiab] OR “Myocardial Infarction”[tiab] OR “Cerebral Infarction”[tiab])))

## Cochrane

(( (Cell Phones OR Mobile Applications OR Actigraphy OR Wearable Devices OR mhealth OR ( smartphone* OR wearable* OR “activity monitor” OR “activity tracker” OR mhealth OR “m-health” OR “mobile health” OR actigraph* OR “wearable technolog*” OR “wearable device*” OR “smart band*” OR “smart watch*” ):ti,ab ) AND (Exercise OR Motor Activity OR Sedentary OR Physical Fitness OR ( “physical activity” OR exercis* OR “physical fitness” OR “activity level*” OR “exercise behavio*” OR “physical exercise” OR sport ):ti,ab ) AND (Randomized Controlled Trial OR “Clinical Trial, Randomized” OR ( “randomized controlled trial” OR RCT OR “randomised controlled trial” ):ti,ab ) AND (Cardiovascular Diseases OR Stroke OR Myocardial Infarction OR Heart Failure OR Cerebral Infarction OR Heart Diseases OR ( CVD OR “Cardiovascular disease” OR “Cardiovascular Disorder” OR Stroke OR “Brain Infarction” OR “Heart Failure” OR “heart disease” OR MI OR “Myocardial Infarction” OR “Cerebral Infarction” ):ti,ab ) ))

## Web of Science

(

ALL=(

(“Cell Phones” OR “Mobile Applications” OR “Actigraphy” OR “Wearable Devices” OR “mhealth” OR “smartphone*” OR “wearable*” OR “activity monitor” OR “activity tracker” OR “m-health” OR “mobile health” OR “actigraph*” OR “wearable technolog*” OR “wearable device*” OR “smart band*” OR “smart watch*” OR “Mobile Phone*” OR “Cell Phone*” OR “Cellular Phone*” OR “Smartphone*” OR “Smart Phone*” OR “Mobile Telephone*” OR “Acceleromet*”

) )

AND

ALL=(

“Exercise” OR “Motor Activity” OR “Sedentary” OR “Physical Fitness” OR “physical activity” OR “exercis*” OR “physical fitness” OR “activity level*” OR “exercise behavio*” OR “physical exercise” OR “sport” OR “activity pattern*” OR “exercise pattern*” OR “physical behavio*” OR “recreational activity”

)

AND

ALL=(

“Randomized Controlled Trial” OR “Clinical Trial, Randomized” OR “randomized controlled trial” OR “RCT” OR “randomised controlled trial”

)

AND

ALL=(

“Cardiovascular Diseases” OR “Stroke” OR “Myocardial Infarction” OR “Heart Failure” OR “Cerebral Infarction” OR “Heart Diseases” OR “CVD” OR “Cardiovascular disease” OR “Cardiovascular Disorder” OR “Brain Infarction” OR “heart disease” OR “MI” OR “Cerebral Infarction”

)

AND ALL=(“Cell Phones” OR “Mobile Applications” OR “Actigraphy” OR “Wearable Devices” OR “mhealth” OR “smartphone*” OR “wearable*” OR “activity monitor” OR “activity tracker” OR “m-health” OR “mobile health” OR “actigraph*” OR “wearable technolog*” OR “wearable device*” OR “smart band*” OR “smart watch*” OR “Mobile Phone*” OR “Cell Phone*” OR “Cellular Phone*” OR “Smartphone*” OR “Smart Phone*” OR “Mobile Telephone*” OR “Acceleromet*” ) )

