## Supplementary material for "Effectiveness of Physical Activity Interventions Utilizing Wearables and Smartphone Applications for Individuals with Cardiovascular Diseases and Stroke: A Systematic Review and Meta-analysis": Tables

Table 1: Characteristics of Included Studies

| **StudyID** | **Mean age (years)/ N (randomized)** | **Intervention Duration** | **FollowUpTime** | **Measurement Type** | **Control** | **Intervention** | **Behavioral Theory** | **Population** |
| --- | --- | --- | --- | --- | --- | --- | --- | --- |
| Mitropoulos2024 | 53.8/30 | 26 weeks | 24 weeks | Objective | Supervised gym-based resistance training and aerobic exercise | Online home based Resistance Training combined with aerobic exercise | [**Goals and planning**: Goal setting (outcome), Action planning], [**Feedback and monitoring**: Feedback on behaviour, Self-monitoring of behaviour, Self-monitoring of outcome(s) of behaviour, Feedback on outcome(s) of behavior], [**Shaping knowledge**: Instruction on how to perform a behavior], [**Repetition and substitution**: Behavioral practice/rehearsal, Graded tasks] | MI patients |
| Varnfield2014 | 55.7/94 | 6 weeks | 24 weeks | Objective | Standard Cardiac Rehabilitation | The CAP-CR platform used a smartphone for health and exercise monitoring, and delivery of motivational and educational materials to participants via text messages and preinstalled audio and video files (including understanding cardiovascular disease (CVD), symptoms and management) | [**Goals and planning**: Goal setting (behavior), Action planning, Review behavior goal(s)], [**Feedback and monitoring**: Feedback on behaviour, Self-monitoring of behaviour, Self-monitoring of outcome(s) of behaviour], [**Shaping knowledge**: Instruction on how to perform a behavior], [Natural consequences: Information about health consequences], [**Associations**: Prompts/cues], [**Regulation**: Reduce negative emotions] | MI patients |
| Yudi2020 | 56.8/168 | 8 weeeks | 8 weeks | Objective | Standard Cardiac Rehabilitation | a multi-faceted interventioned through a smartphone application. It incorporates all core aspects of cardiac  rehabilitation; namely exercise prescription, dynamic  tracking of cardiovascular risk factors, assessment of dietary habits, heart health education, education on secondary prevention pharmacotherapy as well | [**Goals and planning**: Goal setting (behavior)], [**Feedback and monitoring**: Feedback on behaviour, Self-monitoring of behaviour, Self-monitoring of outcome(s) of behaviour], [**Natural consequences**: Information about health consequences], [**Associations**: Prompts/cues], [**Identity**: Framing/reframing] | CHD, Acute coronary syndrome |
| Beckie2024 | 61.2/47 | 12 weeks | 12 weeks | Objective | Standard Cardiac Rehabilitation | App which displayed progress, education videos, interaction with a health coach, peer support | [**Goals and planning**: Goal setting (behavior), Review behavior goal(s)], [**Feedback and monitoring**: Feedback on behaviour, Self-monitoring of behaviour], [**Social support**: Social support (emotional)], [**Shaping knowledge**: Instruction on how to perform a behavior], [**Natural consequences**: Information about health consequences], [**Associations**: Prompts/cues], [**Self-belief**: Verbal persuasion about capability] | women with CHD |
| Bernal-Jiménez2024 | 59.4/128 | 36 weeks | 36 weeks | Subjective | Standard Cardiac Rehabilitation | mHealth intervention through an app on mobile or tablet. App established aims to achieve PA, food consumption, BP, smoking and therapeutic adherence and provided education | **BT**: Phases of change theory; [**Goals and planning**: Goal setting (behavior), Review behavior goal(s)], [**Feedback and monitoring**: Feedback on behaviour, Self-monitoring of behaviour, Self-monitoring of outcome(s) of behaviour], [**Shaping knowledge**: Instruction on how to perform a behavior], [**Natural consequences**: Information about health consequences], [**Associations**: Prompts/cues], [**Repetition and substitution**: Behavioral practice/rehearsal, Habit formation] | Coronary Artery Disease (CAD) |
| Patel2021 | 58/196 |  | 24 weeks | Objective | Individualized, self-set goals and pre-set goals | Gamification: The game included points and levels that were run automatically (participants did not have to actively play the game—just strive for step goals) and provided daily feedback via text message on their progress. | [**Goals and planning**: Goal setting (behavior), Review behavior goal(s)], [**Feedback and monitoring**: Feedback on behaviour, Self-monitoring of behaviour], [**Associations**: Prompts/cues], [**Repetition and substitution**: Graded tasks], [**Reward and threat**: Non-specific reward], [**Scheduled consequences**: Behavior cost] | atherosclerotic cardiovascular disease (ASCVD) or a 10-year ASCVD risk score greater than or equal to 7.5% |
| Willinger2023 | 15.1/97 | 12 weeks | 12 weeks | Objective | Wearable without receiving further instructions | Study participants received brief and informative text and video messages on their smartphone daily over a period of 12- weeks | **BT**: Bandura’s social cognitive; [**Goals and planning**: Review behavior goal(s)], [**Feedback and monitoring**: Self-monitoring of behaviour], [**Shaping knowledge**: Instruction on how to perform a behavior], [**Natural consequences**: Information about health consequences], [**Associations**: Prompts/cues], [**Repetition and substitution**: Behavioral practice/rehearsal], [**Self-belief**: Verbal persuasion about capability] | all patients at an age of 12 to 18 years, with moderate-to-complex CHD |
| Grau-Pellicer2019 | 62.9/34 |  | 12 weeks | Objective | Conventional Rehabilitation | to have bidirectional feedback: participants could visualize results and exchange messages with the researchers | [**Goals and planning**: Goal setting (behavior), Review behavior goal(s)], [**Feedback and monitoring**: Feedback on behaviour, Self-monitoring of behaviour], [**Social support:** Social support (emotional)], [**Shaping knowledge**: Instruction on how to perform a behavior], [**Repetition and substitution**: Behavioral practice/rehearsal, Graded tasks], [**Identity**: Identity associated with changed behavior], [**Self-belief**: Verbal persuasion about capability] | ischemic or hemorrhagic stroke |
| Duscha2018 | 59.9/25 | 12 weeks | 12 weeks | Objective | Usual care | mHealth + Human coach | [**Goals and planning**: Goal setting (behavior), Review behavior goal(s)], [**Feedback and monitoring**: Feedback on behaviour, Self-monitoring of behaviour], [**Social support**: Social support (unspecified)], [**Shaping knowledge**: Instruction on how to perform a behavior], [**Associations**: Prompts/cues], [**Repetition and substitution**: Graded tasks], [**Self-belief**: Verbal persuasion about capability] | CVD patients |
| Xu2022 | 53.7/72 | 12 weeks | 24 weeks | Objective | Personalized daily step goals were set in the WeChat applet backstage based on patients’ baseline daily step counts and the goals increased gradually from the baseline by 15% each week during the first 6 weeks and then remained constant during the last 6 weeks | Patients in the individual group received 140 points every Monday (20 points per day), and if they met their daily step goal, no points were deducted; if they did not, 20 points were deducted. | [**Goals and planning**: Goal setting (behavior), Review behavior goal(s)], [**Feedback and monitoring**: Feedback on behaviour, Self-monitoring of behaviour], [**Social support**: Social support (unspecified), Social support (emotional)], [**Comparison of behaviour**: Social comparison], [**Repetition and substitution**: Graded tasks], [**Reward and threat**: Material reward (behavior), Non-specific reward, Social incentive], [Scheduled consequences: Behavior cost], [**Self-belief**: Verbal persuasion about capability] | CHD (including acute myocardial infarction and unstable angina) |
| Paul2016 | 56.3/23 | 6 weeks | 6 weeks | Objective | Usual care | Individual and group ‘rewards’ for achieving goals were provided. As the participant reached their target number of steps, their fish’s fins and tail grew. If all four members reached their step count target on at least five days of the week then the group was rewarded by another sea creature being added to their fish tank e.g. sea horse or crab | [Goals and planning: Goal setting (behavior), Review behavior goal(s)], [Feedback and monitoring: Feedback on behaviour, Self-monitoring of behaviour], [Social support: Social support (unspecified)], [Comparison of behaviour: Social comparison], [Repetition and substitution: Graded tasks], [Reward and threat: Non-specific reward, Social reward] | Stroke |
| Jo2024 | 57/41 | 6 weeks | 6 weeks | Objective | Standard Cardiac Rehabilitation | The application included exercise modes for warm-up, aerobic, stretching, and resistance exercises, and it provided exercise records and a diary | [**Goals and planning**: Goal setting (behavior), Review behavior goal(s)], [**Feedback and monitoring**: Feedback on behaviour, Self-monitoring of behaviour], [**Shaping knowledge**: Instruction on how to perform a behavior], [**Repetition and substitution**: Behavioral practice/rehearsal] | post-AMI patients |
| Lunde2020 | 59/102 | 54 weeks | 54 weeks | Objective | Usual care | The app provided automatic reminders and evaluations of tasks and weekly goal achievement.The patients received short, tailored, individualized motivational feedback directly through the app 1–3 times a week. Additionally, they received comprehensive individual feedback via email once a week for the first 12 weeks and every fourth week for the rest of the year. | [**Goals and planning**: Goal setting (behavior), Problem solving, Action planning, Review behavior goal(s)], [**Feedback and monitoring**: Feedback on behaviour, Self-monitoring of behaviour], [**Social support**: Social support], [**Shaping knowledge**: Instruction on how to perform a behavior], [**Associations**: Prompts/cues], [**Self-belief**: Verbal persuasion about capability] | CAD and valve surgery |
| Gallagher2023 | 61.2/390 | 32 weeks | 32 weeks | Subjective | Usual care | Gamification centered on a cartoon heart avatar as a virtual representation of the user’s health. Users could maintain their own health by completing the active behaviour change components and, by doing so, earn coins to purchase and provide healthy food, exercise, medications, and relaxation items | [**Feedback and monitoring**: Feedback on behaviour, Self-monitoring of behaviour, Self-monitoring of outcome(s) of behaviour], [**Shaping knowledge**: Instruction on how to perform a behavior], [**Natural consequences**: Information about health consequences], [**Comparison of behaviour**: Social comparison] [**Associations**: Prompts/cues], [**Repetition and substitution**: Behavioral practice/rehearsal, Graded tasks], [**Reward and threat**: Non-specific reward, Social reward], [**Identity**: Framing/reframing], [**Self-belief**: Verbal persuasion about capability] | CHD |

Table 2: Distribution of Behavioral Change Techniques

| BCT Name | Count |
| --- | --- |
| Self-monitoring of behaviour | 14 |
| Feedback on behaviour | 13 |
| Goal setting (behavior) | 11 |
| Review behavior goal(s) | 11 |
| Instruction on how to perform a behavior | 10 |
| Prompts/cues | 9 |
| Graded tasks | 7 |
| Verbal persuasion about capability | 7 |
| Information about health consequences | 6 |
| Behavioral practice/rehearsal | 6 |
| Self-monitoring of outcome(s) of behaviour | 5 |
| Non-specific reward | 4 |
| Social support (unspecified) | 4 |
| Action planning | 3 |
| Social support (emotional) | 3 |
| Social comparison | 3 |
| Framing/reframing | 2 |
| Social reward | 2 |
| Behavior cost | 2 |
| Social incentive | 1 |
| Material reward (behavior) | 1 |
| Goal setting (outcome) | 1 |
| Reduce negative emotions | 1 |
| Identity associated with changed behavior | 1 |
| Habit formation | 1 |
| Feedback on outcome(s) of behavior | 1 |
| Problem solving | 1 |
